# Modelling the Long-Term Effects of Covid-19 Cancer Services Disruption on Patient Outcome in Scotland

**DOI:** 10.1101/2021.01.17.21249993

**Authors:** Jonine D Figueroa, Ewan Gray, Yasuko Maeda, Peter S Hall, Melanie Mackean, Kenneth Elder, Farhat V N Din, Malcolm G Dunlop, David Weller

## Abstract

**Background:** Modelling the long-term effects of disruption of cancer services and minimising any excess cancer mortality due to the Covid-19 pandemic is of great importance. Here we adapted a stage-shift model to inform service planning decisions within NHS Scotland for the ‘‘Detect Cancer Early’ tumours, breast, colorectal and lung cancer which represent 46% of all cancers diagnosed in Scotland.

**Methods & Data:** Lung, colorectal and breast cancer incidence data for years 2017-18 were obtained from Public Health Scotland Cancer Quality Performance Indicators (QPI), to define a baseline scenario. The most current stage-specific 5-year survival data came from 2009-2014 national cancer registry and South East Scotland Cancer Network (SCAN) QPI audit datasets. The Degeling et al., inverse stage-shift model was adapted to estimate changes in stage at diagnosis, excess mortality and life-years lost from delays to diagnosis and treatment due to Covid-19-related health services disruption. Three and 6-month periods of disruption were simulated to demonstrate the model predictions.

**Results:** Approximately, 1-9% reductions in stage I/II presentations leading up to 2-10% increases in stage III/IV presentations are estimated across the three cancer types. A 6-month period of service disruption is predicted to lead to excess deaths at 5 years of 32.5 (31.1, 33.9) per 1000 cases for lung cancer, 16.5 (7.9, 24.3) for colorectal cancer and 31.6 (28.5, 34.4) for breast cancer.

**Conclusions:** Disruption of cancer diagnostic services can lead to significant excess deaths in following years. Increasing diagnostic and capacity for cancer services to deal with the backlog of care are needed. Real time monitoring of incidence and referral patterns over the disruption and post-disruption period to reduce excess deaths including more rapid incidence data by stage and other key tumour/clinical characteristics at presentation for key cancer cases (on a quarterly basis). Real time monitoring in cancer care and referral patterns should help inform what type of interventions are needed to reduce excess mortality and whether different population subgroups require public health messaging campaigns. Specific mitigation measures can be the subject of additional modelling analysis to assess the benefits and inform service planning decision making.

## Introduction

The Covid-19 pandemic has been a public health emergency with widespread direct, and indirect, impacts. It is clear that reducing the infection rate is the first line of defence to maintain health services and the public’s confidence to safely seek care. However, with increasing rates of Covid-19 there is an increasing burden and unfortunate need to prioritise services that will prevent the most deaths arising from disruption to cancer services. Further, what types of public health messaging are needed and whether certain high-risk groups need to be targeted could be informed by routinely collected data. Important decisions need to be made as to the prioritisation in resumption of services, continuing trade-offs between service capacity and Covid-19 safety measures, and further restrictions in the case of increased disruption due to pandemic.

Existing modelling studies ^1-5^provide only limited information on the long-term effects of disruption of cancer services due to the Covid-19 pandemic. Recent calls for real time data to monitor the impacts on mortality and presentation are needed. The impacts will vary for different populations and in this paper, we have modelled the potential mortality impacts for the three common Detect Cancer Early (DCE) (https://www.isdscotland.org/Health-Topics/Cancer/Detect-Cancer-Early/) tumour types in Scotland (lung, colorectal cancer, breast) in order to inform service planning decisions within NHS/Public Health Scotland (PHS). This analysis may be useful if adapted to other settings internationally as many health systems face similar decisions and are keen to determine key metrics that need to be routinely captured in order to monitor the situation and provide mitigation measures to minimize mortality impacts.

This paper reports the model structure, input parameters and results under a selection of scenarios. Lung, colorectal and breast are common cancers and are responsible for a large proportion of cancer mortality and morbidity in Scotland ^6^. We focused on generating predictions of excess deaths and life-years lost, but acknowledge that, while this is a key consideration, this is by no means the only factor relevant to decisions regarding cancer services.

This provides a framework from which more specific analysis can be derived to inform cancer site specific service planning decisions. While this work is focused on service disruptions due to the Covid-19 pandemic, such modelling could be important beyond the pandemic; it has the potential to help with future cancer services planning and also assist in prioritising cancer data intelligence.. Our aims were to: 1) estimate the potential impact of disruption of cancer diagnostic services due to Covid-19 on cancer patient mortality in the medium to long-term 2) explore how modelling of medium to long term outcomes following service disruption can inform service planning decisions. The decisions considered relate to priorities in service resumption as well as potential further restriction in the event of increased pandemic threats.

## Sources of data to populate our model

### Incidence

Data on incidence and stage distribution of cancers in years prior to 2020 was sought to estimate the expected incidence for 2020. We used an average of reported incidence by stage for the years 2017 and 2018 obtained from the PHS (formerly known as information services division -ISD),^6^ DCE ^7^dataset and by SCAN QPI Audit.

### Survival

Stage specific survival data for up to 5 years following diagnosis were obtained from different sources for lung, colorectal and breast cancer.

#### Lung cance

No Scottish data were immediately accessible therefore published data for England were used. Estimates were taken from the Office of National Statistics (ONS) publication “Cancer Survival in England: adults diagnosed between 2013 and 2017 and followed up to 2018” which uses data combined from English regional cancer registries.

#### Colorectal cancer

Data were obtained from an existing extract from the SCAN cancer audit data set. The extract included all incident cases within Lothian health board between January 2009 and December 2014 with follow-up of vital status until at least December 2019.

#### Breast cancer

stage and mode of detection (screening/non-screening) specific survival data were obtained from a pre-existing extract from the Scottish Cancer Registry ^8^.

## Methods

This study adapted the Degeling et al inverse stage-shift model ^1^ to estimate excess mortality and life-years lost from delays to diagnosis and treatment from health services disruption. Adaptation included changes to the model structure as well as input of Scotland specific incidence and survival parameters derived from the above data.

### Inverse stage-shift model

A baseline distribution for the proportion of incident cancers detected at each stage (I-IV and stage unknown) for the period of study was defined based on historical data as detailed above.

Scenarios of service disruption leading to delays in diagnosis are simulated by estimating the stage distribution of incident cases following a period of delay. Changes in stage distribution at diagnosis may be estimated by applying probabilities of stage progression (stage-shift) for fixed periods of delay.

The stage-shift probabilities for a given period of delay are calculated based on the observed relationship between delays in time to treatment initiation (TTI) and survival. The TTI-survival relationship is informative to stage-shift if one assumes that higher mortality following delay arises from stage-shifts, i.e. stage mediates the effect of delay in TTI on survival.

Treatment progression is calculated following the same equations separately for each (j) stage. The expected time to progression to the next stage (*T*_*j*_) is calculated based on the input parameters of hazard ratio for delay in time to treatment initiation (*HR TTI*_*j*_), the corresponding time period of delay that *HR TTI*_*j*_ applies to (*t*_*j*_), and the mortality hazard ratio for stage *j* compared to stage j+ *i* (*HR mort*_*j*_), as follows:

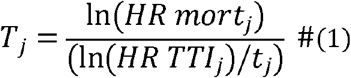

The even rate (*λ*_j_) for progression to stage *j* + 1 is 1/*T*_*j*_. If an exponential distribution of stage progression times is assumed then this event rate can be used to calculate the probability of a stage transition (*P*_*j*_) conditional on the period of delay (*t*_*d*_) as:

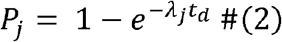

Note that in this method the progression rates and probabilities will change when either the TTI HR or stage-specific survival estimates change. I.e. stage transition probabilities will vary over place and time due to different stage-specific survival estimates even if the same TTI HR estimates are applied.

### Adaptions to the model

Transitions considered were from stage I to stages II-IV and from stage II to stages III-IV. Stage transitions from III to IV were not included as it was assumed that cases in these stages would still present and be diagnosed at approximately normal rates through emergency and urgent referral pathways. Progression to stage III/IV from earlier stages was assumed to be proportional to the current relative proportions of stage III/stage IV incident cases.

Stage transitions were calculated sequentially, i.e. calculate probability of transition from stage I to stage III/IV, then conditional on not transitioning to stage III/IV calculate the probability of transitioning to stage II.

Stage transition probabilities were applied to the baseline stage distribution to calculate the resulting stage distribution following stage shift. Survival for the cohort were estimated by calculating the weighted average of survival across stages weighted by the post stage-shift proportions in each stage. Excess mortality at 5 years was calculated by taking the difference in survival at 5 years compared to the no delay scenario.

TTI HR estimates were obtained from ^9^, these were 1.032 for lung, 1.005 for colorectal and 1.018 for breast. The same TTI HR was used for all stages although the TTI HR could be varied by stage in future analysis. Furthermore, it is possible to include within stage effects on mortality through application of TTI HR to all cases in addition to stage changes if one wished to assume the effect is additive.

### Scenarios used in the analysis

The precise degree of service disruption is difficult to estimate with the relatively premature data currently available. To demonstrate the model predictions given plausible periods of disruption values of 3- and 6-month disruptions were chosen. This can be refined in a future iteration of the modelling analysis as suitable data becomes available.

The base case analysis used the best estimate parameter values listed in table SA1. One-way sensitivity analysis (OWSA) for TTI HR parameters was conducted to demonstrate the how sensitive the excess deaths outcome is to variation in these parameters. Probabilistic sensitivity analysis was used to calculate 95% confidence intervals capturing the uncertainty in the sampling distribution of the parameters for HR TTI parameters. Note this does not reflect other uncertainty such as uncertainty about the appropriate model structure. In this type of modelling exercise unquantified model structural uncertainty may be the most important source of uncertainty. Model and input data can be found at github: https://github.com/EwanGray/cancer_covid_scot

## Results

Table 1 shows the baseline stage distributions predicted for 2020 from prior years data. The scenarios following 3- or 6-month disruptions show the stage distributions following the stage-shifts that are predicted to occur in these scenarios. Stage-shifts lead to a general pattern of upgrading of stage at diagnosis. This varies by cancer site due to differing baseline stage distributions and also different progression rates.

**Table 1.** Predicted stage distributions baseline (no disruption) and following delays of 3 or 6 months

| Scenario | Stage (%) |  |  |  |  |
| --- | --- | --- | --- | --- | --- |
|  | I | II | III | IV | Unknown |
| Lung<br>(N = 4622) |  |  |  |  |  |
| Baseline | 17.9 | 7.44 | 21.6 | 45.8 | 7.22 |
| 3 months | 15.7 | 7.48 | 22.3 | 47.3 | 7.22 |
| 6 months | 11.2 | 6.97 | 23.9 | 50.8 | 7.22 |
| Colorectal<br>(N = 3366) |  |  |  |  |  |
| Baseline | 16.4 | 23.9 | 25 | 23.8 | 10.8 |
| 3 months | 16 | 23.3 | 25.5 | 24.3 | 10.8 |
| 6 months | 14.8 | 21.7 | 27 | 25.7 | 10.8 |
| Breast<br>(N = 4407) |  |  |  |  |  |
| Baseline | 40.4 | 45.9 | 7.54 | 5.11 | 1.07 |
| 3 months | 37.8 | 44.6 | 9.81 | 6.65 | 1.07 |
| 6 months | 31.4 | 41.1 | 15.77 | 10.69 | 1.07 |

Table 2 shows the estimated excess mortality for 1-5 years under 3- and 6-months disruption scenarios. The excess mortality differs by time and cancer type with lung cancer showing the greatest excess mortality early (1 year) and breast cancer later (5 years). Table 3 displays the estimated excess deaths at five years and LY lost over 10 years under 3- and 6-month disruption scenarios. In total for the Scottish population, 48 and 150 excess deaths from lung cancer are estimated for 3 and 6 months of disruption respectively. The corresponding numbers of excess deaths for colorectal cancer are 15 and 55, while for breast cancer they are 29 and 139.

**Table 2.** Predicted excess deaths from 1-5 years due to 3 or 6 month delays by cancer type

| Cancer<br>Duration<br>of<br>disruption | Excess deaths assessed at each time point |  |  |  |  |
| --- | --- | --- | --- | --- | --- |
|  | Year 1 | Year 2 | Year 3 | Year 4 | Year 5 |
| **Lung** |  |  |  |  |  |
| 3 months | 60.6 | 59.5 | 56.4 | 52.4 | 48 |
| 6 months | 192.4 | 188.1 | 177.7 | 164.5 | 150 |
| **Colorectal** |  |  |  |  |  |
| 3 months | 9.52 | 13.5 | 15.2 | 15.2 | 14.7 |
| 6 months | 35.92 | 50.8 | 57.3 | 57.2 | 55.4 |
| **Breast** |  |  |  |  |  |
| 3 months | 7.47 | 22.8 | 29.6 | 35.4 | 38.7 |
| 6 months | 26.95 | 82.3 | 106.5 | 127.4 | 139.1 |

**Table 3.**
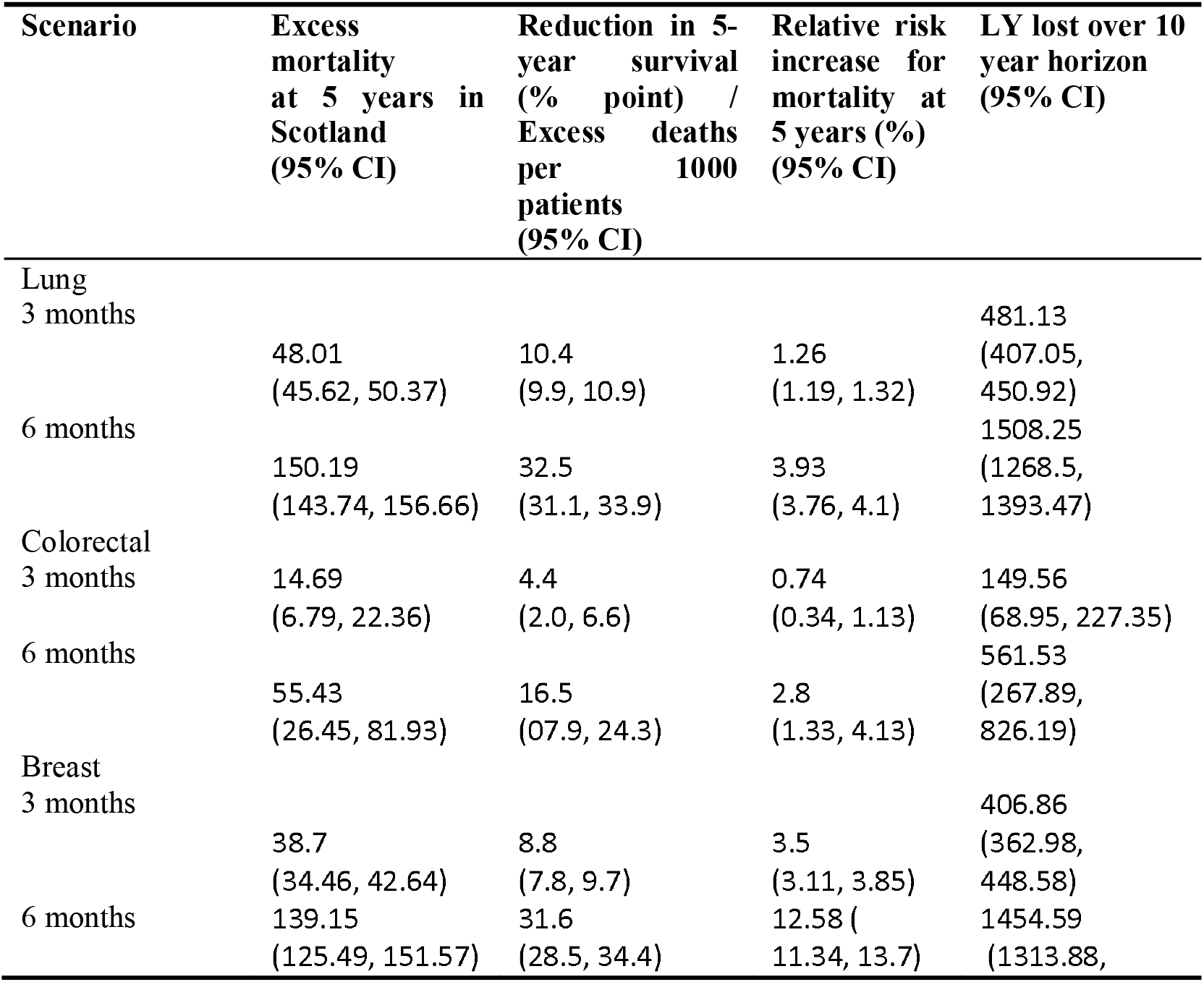
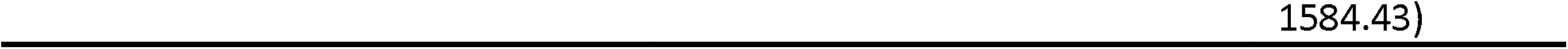
Predicted excess mortality at 5 years and LY lost over 10 years

| Scenario | Excess mortality at 5 years in Scotland (95% CI) | Reduction in 5-year survival (% point) / Excess deaths per 1000 patients (95% CI) | Relative risk increase for mortality at 5 years (%) (95% CI) | LY lost over 10 year horizon (95% CI) |
| --- | --- | --- | --- | --- |
| **Lung** |  |  |  |  |
| 3 months | 48.01<br>(45.62, 50.37) | 10.4<br>(9.9, 10.9) | 1.26<br>(1.19, 1.32) | 481.13<br>(407.05, 450.92) |
| 6 months | 150.19<br>(143.74, 156.66) | 32.5<br>(31.1, 33.9) | 3.93<br>(3.76, 4.1) | 1508.25<br>(1268.5, 1393.47) |
| **Colorectal** |  |  |  |  |
| 3 months | 14.69<br>(6.79, 22.36) | 4.4<br>(2.0, 6.6) | 0.74<br>(0.34, 1.13) | 149.56<br>(68.95, 227.35) |
| 6 months | 55.43<br>(26.45, 81.93) | 16.5<br>(07.9, 24.3) | 2.8<br>(1.33, 4.13) | 561.53<br>(267.89, 826.19) |
| **Breast** |  |  |  |  |
| 3 months | 38.7<br>(34.46, 42.64) | 8.8<br>(7.8, 9.7) | 3.5<br>(3.11, 3.85) | 406.86<br>(362.98, 448.58) |
| 6 months | 139.15<br>(125.49, 151.57) | 31.6<br>(28.5, 34.4) | 12.58<br>(11.34, 13.7) | 1454.59<br>(1313.88, |

OWSA for TTI HR demonstrate that this parameter is highly influential in determining the predicted number of excess deaths as can be seen in Figure 1. This parameter is set based on estimates from regression analysis using observational data which has significant potential bias from residual confounding. A range of plausible values could be considered when thinking about the scope of potential impact.

**Figure 1:**
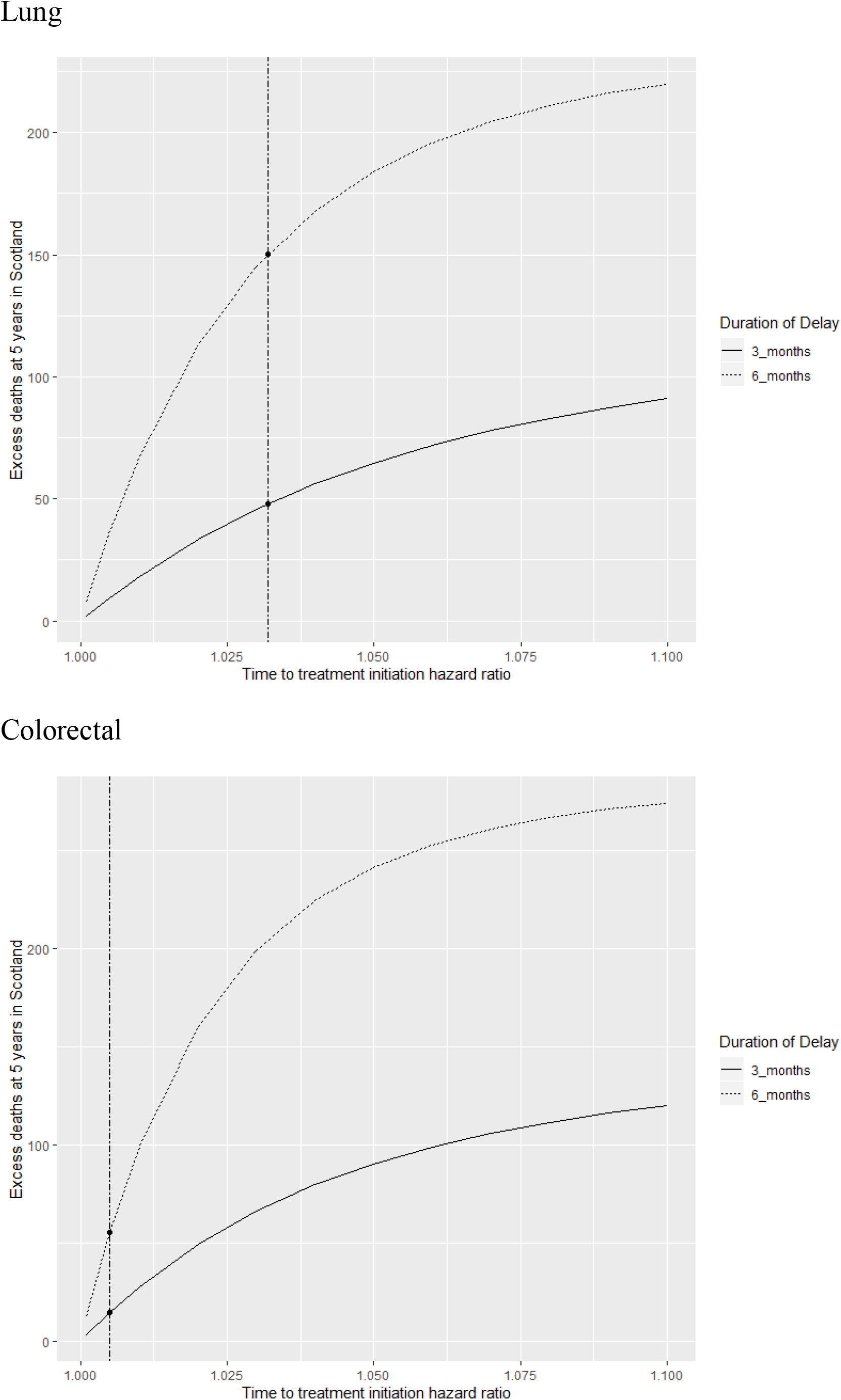
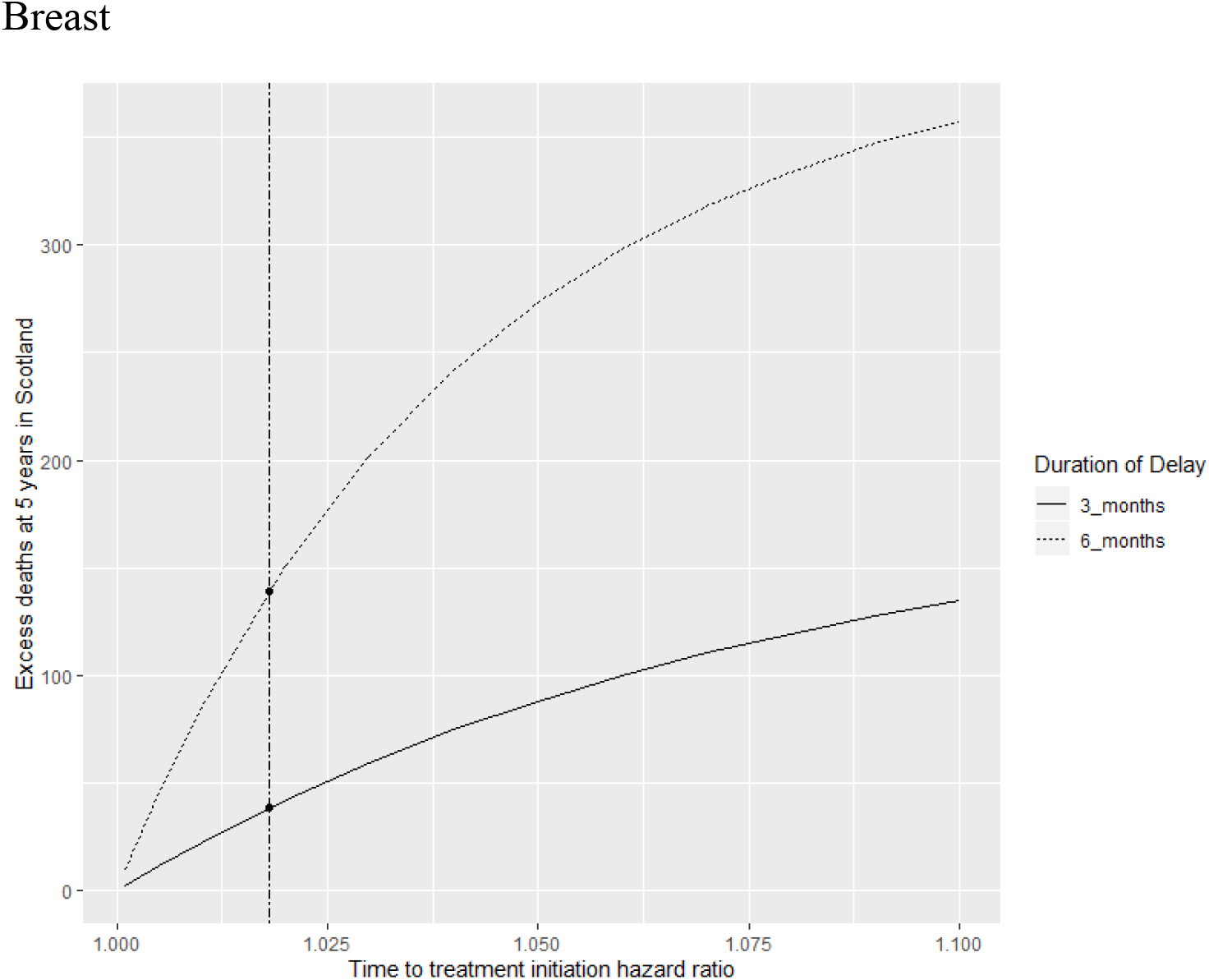
Sensitivity analysis varying time to treatment initiation hazard ratio parameter. Horizontal dashed line indicates the HR used in the modelled scenarios for each cancer.

## Discussion

It is clear that disruption of cancer services at diagnosis, treatment and screening can lead to significant excess deaths and our analysis using Scottish data show, using a model of cancer stage progression that these excesses differ by type of cancer and length of disruption. Keeping Covid-19 rates of infection is, of course, imperative in reducing loss of life. However, we now know that mitigating against the indirect impacts of the epidemic is also extremely important – minimising Covid-19-related disruption to cancer screening, diagnostic and treatment services is an important example.

Specific mitigation measures can be the subject of additional modelling analysis to assess the benefits and inform service planning decision making. For example, we have recently reported the relative importance of screening and symptomatic route of presentation in breast cancer and it may be important for screening services to prioritise those patients that are at highest risk to provide the most benefit^10-12^. For colorectal cancers whose diagnostic procedures are more invasive and require considerable patient preparation time, colonoscopy disruption and use of screening tests to select patients that are most likely to have cancer remain uncertain. From this pandemic, cancer services need to think creatively about strategies to provide new diagnostic pathways due to strained services and to understand trade-off for more stringent safety in relation to Covid-19.

While data to support whether a stage shift will take some years, early indicators that could be monitored with routine data of stage at presentation would be reduced incidence of early-stage cancers, particularly for breast and colorectal cancers where screening has been shown to improve earlier diagnosis and lower stage at presentation. While stage is a crude measure, future analysis should also evaluate whether outcomes are poorer such as 30-60 day mortality measures compared to usual services. We will be continuing monitoring these trends to determine if the resumption of paused services improves early presentation or not. Such data will also assist to quantify the potential benefits of public health efforts to encourage patients to seek healthcare, which may have already impacted breast cancer patients as we see little difference compared to 2019 in number of non-screened referrals.

Our analysis using a stage shift model was similar to other models recently presented focusing on English data ^13^. Mortality impact of delays to treatment varies by cancer site and timing of excesses with lung having more excesses early (1 year), Colorectal cancer midway (3 Year) and breast later (5 years). Hence, such data support focusing efforts to minimise excess by cancer site for the short, medium- and long-term planning of cancer services.

Real time data will be monitored to track patterns for the DCE in terms of referral patterns. In particular, we anecdotally see some changes in our local NHS regional data increased GP referrals for breast cancer that might have compensated for the paused screening service and hence may result in fewer excess deaths. On the other extreme, data suggest many fewer lung cancer cases which has been reported in other studies. Routine NHS data should be more regularly monitored and reported on as these can be important objective indicators of whether public health campaigns, mitigation measures, or patient behaviours are changing that would cumulatively impact stage shifts.

Strengths are data access on survival by stage and incidence to model the Scottish situation including early data on stage at diagnosis from April diagnosed cases due to two-month lag using first treatment date as the reference point. Limitations are these data represent early days and model assumptions on the time to treatment impact but future efforts with data will improve such modelling exercises for improved efforts to inform planning of cancer services.

In conclusion, we endorse current efforts urging patients not to wait if they experience symptoms. The ‘Getting the right care in the right place’ campaign launched 14 July 2020 perhaps may have helped improve referrals and data to determine if indeed it did will be forthcoming. For now, making sure patients don’t delay in seeking help about symptoms of potential oncological significance and to get their cancers detected earlier when treatments and surgeries can be most effective.

## Data Availability

Model and input paramaters are available on github

https://github.com/EwanGray/cancer_covid_scot

## Acknowledgement

We thank the NHS staff who collected the data and specifically Lorna Bruce, Jude Chalmers, Christina Lilley, Maria McMenemy, Alistair Smith and David Walker from NHS Lothian/SCAN for input and comments.

### Appendix

**Table SA 1.** Input parameters

| Parameter | Value(s) |  |  |  |  |  |  |
| --- | --- | --- | --- | --- | --- | --- | --- |
| HR TTI |  |  |  |  |  |  |  |
| HR TTI - lung | 1.032 |  |  |  |  |  |  |
| HR TTI - colorectal | 1.005 |  |  |  |  |  |  |
| HR TTI - breast | 1.018 |  |  |  |  |  |  |
| Annual Incidence |  |  |  |  |  |  |  |
| Lung | 4622 |  |  |  |  |  |  |
| Colorectal | 3366 |  |  |  |  |  |  |
| Breast | 4407 |  |  |  |  |  |  |
| Baseline stage distribution |  |  |  |  |  |  |  |
| Lung | I | II | III | IV | unknown |  |  |
|  | 17.9 | 7.44 | 21.6 | 45.8 | 7.22 |  |  |
| Colorectal | I | II | III | IV | unknown |  |  |
|  | 16.4 | 23.9 | 25 | 23.8 | 10.8 |  |  |
| Breast | I | II | III | IV | unknown |  |  |
|  | 40.4 | 45.9 | 7.54 | 5.11 | 1.07 |  |  |
| Baseline 5-year Survival |  |  |  |  |  |  |  |
| Lung | Stage | 0 | 1 | 2 | 3 | 4 | 5 |
|  | I | 1 | 0.852 | 0.752 | 0.663 | 0.585 | 0.516 |
|  | II | 1 | 0.687 | 0.561 | 0.459 | 0.375 | 0.306 |
|  | III | 1 | 0.46 | 0.325 | 0.23 | 0.163 | 0.115 |
|  | IV | 1 | 0.177 | 0.11 | 0.068 | 0.042 | 0.026 |
|  | U | 1 | 0.519 | 0.453 | 0.395 | 0.344 | 0.3 |
| Colorectal | Stage | 0 | 1 | 2 | 3 | 4 | 5 |
|  | I | 1 | 0.979 | 0.95 | 0.924 | 0.876 | 0.844 |
|  | II | 1 | 0.937 | 0.892 | 0.845 | 0.792 | 0.739 |
|  | III | 1 | 0.902 | 0.807 | 0.723 | 0.664 | 0.619 |
|  | IV | 1 | 0.451 | 0.24 | 0.149 | 0.109 | 0.093 |
|  | U | 1 | 0.722 | 0.58 | 0.49 | 0.427 | 0.391 |
| Breast | Stage | 0 | 1 | 2 | 3 | 4 | 5 |
|  | I | 1 | 0.995 | 0.986 | 0.973 | 0.959 | 0.943 |
|  | II | 1 | 0.989 | 0.964 | 0.935 | 0.906 | 0.879 |
|  | III | 1 | 0.962 | 0.88 | 0.816 | 0.769 | 0.721 |
|  | IV | 1 | 0.929 | 0.789 | 0.739 | 0.676 | 0.65 |
|  | U | 1 | 0.994 | 0.973 | 0.938 | 0.899 | 0.845 |

| Study | Method | Main Prediction | Key limitations of method |
| --- | --- | --- | --- |
| Lai et al [Preprint] | Estimated one year mortality for 24 cancer sites (types) [and 15 non-cancer 'comorbidity clusters']. The effect of disruption was calculated based on "relative impact of the emergency" (multiplier to mortality rate) and proportions of the population affected by the emergency. A range of values for relative impact and proportion of population affected were used to produce tabulated results. Plausible ranges are discussed. | "Under conservative assumptions of the emergency affecting only people with newly diagnosed cancer (incident cases) at COVID- 19 PAE of 40%, and an RIE of 1.5, the model estimated 6,270 excess deaths at 1 year in England" | Used a range of estimates of the relative impact of disruption on survival. Therefore, no point estimate or probability distribution of impacts is presented.<br>The mortality impact is not connected to any mechanism, it is purely an assumed increase to mortality.<br>[Compared to the studies below this study assumes both degree of disruption and mortality effect, while those below only assume degree of disruption and mechanism of mortality effect from disruption.] |
| Sud et al [Preprint] | Hazard ratios for per day delay in time to treatment, applied to age, stage-specific cancer survival estimates. Hazard ratios from prior observational studies and baseline from 2013-17 England incidence & 5-year survival. | "six months would cause attributable death of 10,555 of these individuals with loss of 205,024 life years" | Hazard ratios for the effect of a day of delay in treatment initiation are obtained from observational data. Regression strategies to adjust for possible confounding factors are quite limited (difficult to fully capture how delayed cases differ from non-delayed cases) therefore there is substantial potential for bias from residual confounding. |
| Degeling et al [Preprint] | Stage shift model for breast, CRC, lung cancer and melanoma. Stage shift probabilities over a period of delay are inferred from the ratio of hazard ratios (for overall survival) per week delay vs stage I to stage II. Melanoma estimated differently from window study estimates. Stage-specific 5-year survival applied to | CRC: 11 excess deaths after 5 years and a total of 96 life years lost over a 10-year time horizon.<br>Breast: 25 excess deaths over 5 years, 239 LY over 10 years.<br>Lung: 43 excess deaths over 5 years, 373 LY over 10 years.<br>Melanoma: 270 excess deaths over 5 years, 2584 LY over 10 years. | Assumes that expected survival conditional on stage at detection remains constant before, during and following disruption. Stage progression probability is calculated from estimates of hazard ratios for the effect of time-to-treatment initiation on survival. These underlying estimates are vulnerable |
|  | stage distributions with and without delays. Note: Only applied transitions from stage I to stage II. |  | to bias from residual confounding. |
| Maringe et al [Lancet Oncology] | Excess mortality estimated via shift in route-of-presentation for breast, CRC, lung and oesophageal cancers. 3 scenarios of redistribution of cases from routine diagnosis to urgent 2-week referral and emergency presentation. Route-specific 5-year survival from England 2010 (2012 for lung) cohort of cases. Same data used for incidence. Estimates are stratified by age and excess mortality and life years estimates using life tables with added cancer-specific mortality. | “Across the three different scenarios, compared with pre-pandemic figures, we estimate a 7.9–9.6% increase in the number of deaths due to breast cancer up to year 5 after diagnosis, corresponding to between 281 (95% CI 266–295) and 344 (329–358) additional deaths. For colorectal cancer, we estimate 1445 (1392–1591) to 1563 (1534–1592) additional deaths, a 15.3–16.6% increase; for lung cancer, 1235 (1220–1254) to 1372 (1343–1401) additional deaths, a 4.8–5.3% increase; and for oesophageal cancer, 330 (324–335) to 342 (336–348) additional deaths, 5.8–6.0% increase up to 5 years after diagnosis.” | Assumes that expected survival conditional on mode of presentation remains constant before and during disruption. i.e. Cases that cannot present through routine care instead present as urgent referral or emergency AND have the same survival as those presenting through these routes pre-Covid. This is vulnerable to bias if displaced cases have more indolent biology and therefore better prognosis post-detection regardless of mode of detection. Does not make explicit assumed underlying disease progression (tumour growth, stage upgrade, etc). The method can be thought of as using mode of presentation a proxy for disease progression. Additional arbitrary assumption of 25% of cases affected only for screen-detected breast cancer. This is to adjust for possible overdiagnosis. |

